# Sexual experiences among early adolescents aged 12-14 Years in four districts of Rwanda: a cross-sectional study

**DOI:** 10.1101/2024.02.04.24302317

**Authors:** Valens Mbarushimana, Susan Goldstein, Daphney Nozizwe Conco

## Abstract

**Background:** Globally, early adolescents (10-14 years) represent 8% of the world population, and Africa accounts for 25% of them. Although a minority of early adolescents have initiated sexual intercourse, their sexual curiosity results in the exploration and understanding of sexuality. Early sexual intercourse may lead to sexually transmitted infections, HIV/AIDS, early pregnancy or fatherhood, and early marriage. Early sexual activity is associated with high rates of unplanned pregnancy, multiple sexual partners, and other forms of risky sexual behaviours. Understanding sexual activity among early adolescents can contribute to designing interventions that adequately address their needs. However, there is limited information about early adolescents’ sexual activity and the social-ecological factors associated with their sexual experiences. This study aimed to determine the prevalence of sexual activity and the social-ecological factors associated with sexual experiences among early adolescents (12-14 years) in Rwanda.

**Methods:** We conducted a cross-sectional study among early adolescents (12-14 years) from four districts and 16 secondary schools between November and December 2020. A multistage sampling technique was used to select 56 participants from each school, including 28 males and females from grades one and two, who were randomly selected. We used an adapted version of the Illustrative Questionnaire for Interview - Surveys with Young People. Questions focused on nonpenetrative and penetrative sexual experiences in addition to sociodemographic and other social-ecological characteristics. Ethical clearance was obtained from the University of Rwanda and the University of the Witwatersrand, Johannesburg. Written parental or legal guardian consent and participants’ assent were obtained. We conducted the data analysis in Stata 14.2 and used descriptive statistics (frequencies and proportions) and bivariate and multivariate logistic regression analyses with 95% confidence intervals (CIs) and a significance level of p-value <0.05.

**Results:** The study included 811 participants, 55.1% of whom were aged 14, 30.5% were aged 13, and 14.4% were aged 12. Most participants (n=539, 73.5%) lived with both parents, and 48.8% (n=395) described the socioeconomic status of their households as well-off. Nearly 81% (n=658) of the participants indicated that they had experienced non-penetrative sex, and 53 of 759 participants (7%) reported that they had experienced penetrative sex. The social-ecological factors significantly associated with nonpenetrative sexual experiences were attending parties (AOR=6.8, 95% CI= 1.6-29.2), internet use (AOR=1.9, 95% CI= 1.1-3.3) (individual level), and their fathers’ low education (primary: AOR=2.6, 95% CI= 1.4-5.0; secondary: AOR=1.9, 95%CI: 1.0-3.8) (family level). Individual level factors such as male sex (AOR: 4.6, 95% CI= 1.8-12.4), alcohol consumption (AOR=3.5, 95% CI= 1.4-8.8), watching pornography (3-4 times: AOR=7.5, 95% CI= 1.6-34.9, ≥ five times: AOR= 5.1, 95% CI= 1.7-15.0), being a double orphan (AOR:17.8, 95% CI= 1.9-170.2), discussing sex matters often with one’s father (AOR=6.5, 95% CI= 1.8-23.2) (family and relationship level), and forced sexual intercourse (AOR=8.2, 95% CI= 2.7-25.4) (community level) were social-ecological factors associated with penetrative sexual experience.

**Conclusion:** Nonpenetrative sexual experience was common, with few participants reporting penetrative sexual experience. The social-ecological factors associated with sexual experiences among early adolescents are modifiable and can assist in planning healthy sexual interventions for this age group.

## Introduction

Globally, the population of early adolescents (10-14 years) represents 8% of the world population, and the majority of these individuals live in developing countries [1]. Of the estimated 545 million early adolescents in developing countries in 2016, 143 million (26%) lived in Africa [1]. This population of early adolescents in the developing world will grow by 5% by 2030, mainly in Africa, where it is predicted to grow by 34%; this population will stabilise in Asia and decrease in Latin America and the Caribbean by 2030 [1]. In Rwanda, the fourth population and Housing Census 2012 estimated early adolescents (10-14 years) to constitute 12% of the total population [2]. Sexual experiences during early adolescence are a particular focus of this study because behaviours adopted during this period tend to persist into late adolescence and adulthood; hence, there is a need for timely intervention [3].

Adolescence is a critical period of life during which people undergo significant biological, psychological, and social changes [4], and this period marks the transition from childhood to adulthood, during which major sexual developments occur [5, 6]. The literature categorises adolescence into early (at times called very young) adolescence (10-14 years) and late (15-19 years) adolescence [7–9]. One of the salient changes in early adolescence is the body changes and sexual maturation [1]. As these changes occur in early adolescents, they acquire information, develop attitudes, adopt new behaviours, and initiate sexual relationships [10]. This period provides an opportunity to address sexual violence and gender discriminatory norms before they become solid by late adolescence and adulthood [11]

Although a minority of early adolescents have initiated sexual intercourse, sexual curiosity increases during adolescence and leads to the exploration and understanding of sexuality [5, 10, 12]. Sexual exposure becomes important during this period and may result in sexually transmitted infections, HIV/AIDS, adolescent pregnancy, adolescent fatherhood, and early marriage [5, 10], all of which can have further consequences [13]. In particular, sexual activity in adolescents aged under 15 years is associated with high rates of unplanned pregnancy, multiple sexual partners, and other forms of risky sexual behaviours [14]. Therefore, parents, policymakers and stakeholders must intervene before early adolescents face these risks and understand their behaviours to undertake timely, appropriate interventions.

The literature indicates that the sexual and reproductive health and rights of early adolescents are influenced by their social-ecological conditions [15–18]. Deriving from Bronfenbrenner’s Ecology of Human Development [19], Svanemyr et al. suggest that a social-ecological framework recognises multiple influences on sexual behaviours and health outcomes operating at intrapersonal (e.g., sex, religious affiliation, etc.), interpersonal (e.g., relationships with parents), organisational (e.g., structures such as type of school), community (e.g., access to media) and public policy (e.g., current policies and laws on adolescent sexual and reproductive health) levels [16, 18]. Furthermore, this framework suggests that such influence interacts across these levels and requires focusing on specific behaviours and outcomes (e.g., sexual experience) [16]. Finally, the framework suggests that effective interventions should be conducted at multiple levels [16]. The ecological model is applied to understand the predictors of a wide range of behaviours and sexual health outcomes [16], including sexual experiences among early adolescents. In this study, we reasoned that the nonpenetrative and penetrative sexual experiences of early adolescents occur in this social-ecological environment and that the factors associated with such experience are embedded in this framework.

The literature focusing on early adolescents (10-14) is relatively sparse compared with that on older adolescents (15-19 years) [1, 20, 21]. Most often, surveys on sexual behaviours miss this age group because of their young age, and subsequently, national programmes do not reach them [22]. In addition, many studies emphasise vaginal experience, and a few of them discuss nonvaginal experience during adolescence [5]. Furthermore, the literature conceptualises sexual experience as penile-vaginal intercourse [23]. Nevertheless, sexual development is multidimensional, and credit should also be given to other romantic relationships, such as boyfriend or girlfriend, nonpenetrative sexual experiences including holding, hugging, kissing or touching, and penetrative experiences (vaginal, oral, anal) [23]. However, evidence on the nonpenetrative and penetrative sexual activities of early adolescents is still scant, especially in sub-Saharan Africa. The purpose of this study was to assess the prevalence of all sexual activity among early adolescents (12-14 years) and to determine, from the social-ecological perspective, the factors associated with their sexual experiences in four districts of Rwanda.

## Materials and methods

### Study design

We conducted a quantitative cross-sectional study between December 2019 and December 2020 as part of a larger study of early adolescents’ knowledge, beliefs, and behaviours regarding gender and sexuality: implications for sexual experience and health outcomes in Rwanda.

### Study setting

This study was conducted in 16 secondary schools in four districts with a higher prevalence of adolescent pregnancy based on the data from the Demographic and Health Survey conducted in 2014-2015 [24]. These included two urban districts (Gasabo with 12.5% and Nyarugenge with 8.8%) and two rural districts (Rwamagana: 11.9% and Ruhango: 8.2%) [24].

### Study population and sampling

The study population comprised school-going early adolescents (12-14 years) in the four selected districts. We used a multistage sampling strategy to recruit the study participants from 16 secondary schools. We randomly sampled two public and two private secondary schools in each district. In each school, we selected early adolescents in senior one and senior two classes of lower secondary education levels because, according to the Rwanda curriculum framework, we expected early adolescents (12-14 years) to be in these grades [25]. We randomly sampled one class for each school grade from several other classes and chose the existing class for schools with only one grade. We used a lottery method in each class to randomly select an equal number of boys and girls per grade. Therefore, 14 boys and 14 girls were selected to obtain 28 early adolescents for each grade (senior one or senior two). Consequently, 56 early adolescents were sampled from each school. We collected data from 896 learners from 16 schools but excluded data from 85 learners because they were older than 14 years. The final sample size was 811 early adolescents. All participants provided informed consent from their parents or legal guardians and signed informed assent forms.

### Data collection procedures

We contacted Heads of schools to whom we presented the study approvals and authorisation documentation. This documentation included ethical approvals and other administrative authorisations from the relevant institutions controlling research in Rwanda. We then discussed and set together the data collection plans with Heads of schools and affiliated school staff, including the school counsellors. A research team comprising the principal investigator and eight trained research assistants collected the data. On agreed dates, the Heads of schools and other staff members assisted the research team in connecting with the study participants and facilitated the data collection process. Participants received information about the study’s objective and topics addressed in the questionnaire. Participants were advised that their participation was voluntary and that they had the right to stop answering survey questions whenever they wished. The data were collected in classrooms where participants gathered at adequate distances to keep their answers confidential. The survey questions were read aloud one by one by research assistants, and the participants were given adequate time to record their answers on the questionnaire. Each participant received a hard copy of the survey questionnaire and individual assistance from the research team members to answer the questions. Research team members monitored the entire process of data collection until completion.

### Data collection tool

We adapted and used the World Health Organisation (WHO)’s illustrative questionnaire for interview – survey with young people [26]. This questionnaire was designed to investigate the sexual and reproductive health of young people, including early adolescents [26], and was approved by the WHO [27]. This questionnaire has been used in low- and middle-income countries where high validity has been reported [28], and it has been previously used in studies of older [29–32] and early adolescents [28, 33, 34]. The questionnaire content was supplemented by literature on sexual experiences [1, 13, 23, 35–43]. Professionals translated the questionnaire into the local language (Kinyarwanda) and back-translated it into English to ensure consistency. The adapted version was pretested on 20 early adolescents from two schools who were not part of the sample to ascertain the clarity and conciseness of the questions.

### Measures

#### Outcome variables

In this study, sexual experience comprises nonpenetrative and penetrative/coital sexual experiences. We asked participants if they had ever had a partner using a dichotomous question (yes/no): do you have or have you ever had a girlfriend, boyfriend or partner (someone whom you ’dated’ (dating: going out together unaccompanied by other adults)? The participants who responded (yes) responded to a series of dichotomous questions (yes/no) as indicated in Table 1 to reflect on their involvement in the nonpenetrative activity.

**Table 1:** Measures of participants’ lifetime sexual experience.

| Type of experience | Questions asked | Level of measurement |
| --- | --- | --- |
| Nonpenetrative sexual experience | Did you and your boy/girlfriend/partner have any physical contact, such as hugging?<br>Did you and your boy/girlfriend/partner have physical contact, such as holding hands?<br>Did you and your boy/girlfriend/partner have any physical contact, such as caressing each other's bodies?<br>Did you ever kiss your boy/girlfriend/partner on the lips?<br>Did you ever touch your boy/girlfriend/partner's vagina/penis with your hand?<br>Did you ever stroke your boy/girlfriend/partner's vagina/penis so that she/he climaxed?<br>Did your boy/girlfriend/partner ever touch your penis/vagina with her /his hand?<br>Did your boy/girlfriend/partner ever stroke your penis/vagina so that you climaxed? | Yes/no |
| Penetrative sexual experience | Have you ever had oral sex? (oral sex is when a partner puts his/her mouth on your sex organs or you put your mouth on his/her sex organs)<br>Have you ever had anal intercourse? (when a man inserts his penis into his partner's anus)<br>Have you ever had vaginal intercourse with anyone? (Vaginal intercourse is when a boy/man inserts his penis into a girl's/woman's vagina) | Yes/no |

Lifetime nonpenetrative sexual experience was a composite measure obtained from eight questions on various sexual activities, including hugging, holding hands, caressing, kissing, touching, or stroking genitals, as depicted in Table 1. A positive response (yes) to these activities was considered to indicate the presence of nonpenetrative sexual experience.

Lifetime penetrative sexual experience was also a composite measure generated from three questions on experience of oral sex and anal and/or vaginal intercourse. A positive response (yes) to any of these questions was considered as an indication of lifetime penetrative sexual experience. Because we expected few cases of penetrative sexual experience among early adolescents, we preferred lifetime experience to a past year’s experience for low-frequency sexual behaviours [23, 35].

### Social-ecological covariates

We collected social-ecological covariates at the individual, family relationships, and community levels. At the individual level, we included sex (female/male), age in years (12-14), school grade (senior one or two), and religious affiliation (catholic/protestant/others). We also included information such as ever using a mobile phone (yes/no), ever attending parties where young people danced (yes/no), ever drinking alcohol (yes/no), ever smoking cigarettes (yes/no), ever using of cannabis (yes/no), ever using the internet (yes/no), ever searching for pregnancy or puberty information on the internet (yes/no), and ever watching pornography (never, 1-2 times, 3-4 times, ≥5 times). Covariates included at the family and relationships level were parents’ life status (none, one, both); living with these parents (none, one, both); household economic status as measured in the Ubudehe category (categories one to two and categories three to four) [44], father’s and mother’s level of education (primary or lower, technical and vocational education training-TVET, secondary, higher); ever discussed sex matters with the father, mother, brothers or sisters (often, occasionally, never); and perceived ease in communicating with the father, mother, and brothers or sisters (very easy, average, very difficult, do not see him/her/them). Social-ecological covariates at the community level included access to SRHR information (yes/no), type of school (public/private), ever being forced to have sexual intercourse (yes/no), and ever being touched on private parts (e.g., breast) unwillingly.

### Data analysis

We conducted the data analysis in Stata 14.2 for Windows. We constructed frequency tables for all the sociodemographic characteristics of the participants. We conducted bivariate and logistic regression analyses to assess the factors associated with nonpenetrative and penetrative experiences. All variables with statistically significant results (p<0.05) at the bivariate analysis level were included in the logistic regression analysis, where we assessed the effect of each social-ecological covariate on participants’ sexual experience. We report crude and adjusted odds ratios (ORs) with their 95% confidence intervals (CIs). The results with p<0.05 were considered as significant.

### Ethical considerations

This study received ethical approval from the Institutional Review Board (IRB) of the College of Medicine and Health Sciences at the University of Rwanda (UR-CMHS) (No. 543/CMHS IRB/2019) and the Human Research Ethics Committee (HREC)-Medical of the University of the Witwatersrand (clearance certificate no. M190303). In addition, the National Council for Science and Technology (NCST) in Rwanda issued a research permit no. NCST/482/136/2019 for the current study. Before the data collection, we obtained written informed consent from the participant’s parents or legal guardians and participants’ written informed assent. We collected the data anonymously using personal identification codes on the questionnaires. There was adequate distance between participants for their privacy and data confidentiality reasons. We conducted this study following all national regulations and with the principles governing research involving human subjects and school counsellors were on standby to assist in cases of child distress.

## Results

### Participants’ characteristics

#### Individual characteristics

Of the 811 participants included in the analysis, 413 (50.9%) were girls, and the majority 447 (55.1%) were 14 years old. The overall mean age was 13.4±0.7 years. Most participants, 423 (52.2%), were in senior one. Of the 803 participants who indicated their religious affiliation, 290 (36.1%) participants were affiliated with the Roman Catholic Church, and the majority, 370 (46.1%), were affiliated with the Protestant Church (ADEPR and Adventist). More boys (36.4%) than girls (31.2%), n=788, reported that they had ever used mobile phones.

Almost 11% (n=86) of the 782 participants had ever attended parties where young people danced, including 48 boys (6.1%) and 38 girls (4.8%). Nearly 13% (n=97) of the 776 participants reported ever drinking alcohol, with a significantly greater proportion of boys (7.6%) than girls (4.9%). Furthermore, only 18 of the 788 participants (2.3%) reported ever smoking a cigarette, and 19 participants (2.5%), including 16 boys (4%), indicated having ever used cannabis. In total, 390 of the 762 participants (51.2%) had ever used the internet, with a significantly greater proportion of boys (28.4%) than girls (22.8%). However, 225 of the 620 participants (36.3%) searched SRH information online. Concerning watching pornography, 544 of the 783 participants (69.5%) noted that they had never watched pornographic films showing sexual activity. However, 159 of these participants (20.3%) watched these films 1-2 times, 58 of them (7.4%) have watched these films five times or more, and higher proportions of boys than girls watched such films.

#### Family and relationships

Of the 802 participants who provided the life status of their parents, 720 (89.8%) indicated that both of their parents were alive. Nearly 74% (n=539) of the 733 participants stayed with both parents, while 32 (4.4%) stayed with their parents. The findings indicated that households of 395 participants (48.7%) belonged to the third or fourth category of socioeconomic status, and 35% (n=286) of households belonged to the first or second category (low income). In contrast, 49% of the participants were from high-income households (n=395). However, 130 participants (16%) needed to know the socioeconomic status of their households (Table 2). Primary and lower were the most common education levels for the participants’ fathers (38.2%) and mothers (44%) of the 625 and 693 participants (respectively), followed by secondary education (28.9%) and (27.4%), respectively.

**Table 2:** Distribution of sociodemographic and behavioural characteristics of participants by sex.

|  |  | Male |  | Female |  | All |  |
| --- | --- | --- | --- | --- | --- | --- | --- |
| Characteristics | Categories | n | % | n | % | n | % |
| **Individual characteristics and behaviours** |  |  |  |  |  |  |  |
| Age in years ($n=811$) | 12 | 57 | 14.32 | 60 | 14.53 | 117 | 14.43 |
|  | 13 | 105 | 26.38 | 142 | 34.38 | 247 | 30.46 |
|  | 14 | 236 | 59.30 | 211 | 51.09 | 447 | 55.12 |
| Characteristics | Categories | n | % | n | % | n | % |
|  | Mean | 13.5±0.7 |  | 13.4±0.7 |  | 13.4±0.7 |  |
| School grade (n=811) | Senior 1 | 212 | 53.27 | 211 | 51.09 | 423 | 52.16 |
|  | Senior 2 | 186 | 46.73 | 202 | 48.91 | 388 | 47.84 |
| Religion (n=803) | Catholic | 140 | 35.44 | 150 | 36.76 | 290 | 36.11 |
|  | Protestant | 185 | 46.84 | 185 | 45.34 | 370 | 46.08 |
|  | Others | 70 | 17.72 | 73 | 17.89 | 143 | 17.81 |
| **Behaviours** |  |  |  |  |  |  |  |
| Ever used a mobile phone<br>(n=788) *** | Yes | 287 | 74.35 | 246 | 61.19 | 533 | 67.64 |
|  | No | 99 | 25.65 | 156 | 38.81 | 255 | 32.36 |
| Ever attended parties where<br>young people dance (n=782) | Yes | 48 | 12.24 | 38 | 9.50 | 86 | 10.86 |
|  | No | 344 | 87.76 | 362 | 90.50 | 706 | 89.14 |
| Ever drink alcohol (n=776) * | Yes | 59 | 15.28 | 38 | 9.74 | 97 | 12.50 |
|  | No | 327 | 84.72 | 352 | 90.26 | 679 | 87.50 |
| Ever smoked cigarette (n=788) | Yes | 12 | 3.13 | 6 | 1.49 | 18 | 2.28 |
|  | No | 372 | 96.88 | 398 | 98.51 | 770 | 97.72 |
| Ever use of cannabis (n=775) ** | Yes | 16 | 4.21 | 3 | 0.76 | 19 | 2.45 |
|  | No | 364 | 95.79 | 392 | 99.24 | 756 | 97.55 |
| Ever used internet (n=762) *** | Yes | 216 | 57.60 | 174 | 44.96 | 390 | 51.18 |
|  | No | 159 | 42.40 | 213 | 55.04 | 372 | 48.82 |
| Ever searched SRH information<br>from the internet (n=620) | Yes | 116 | 36.48 | 109 | 36.09 | 225 | 36.29 |
|  | No | 202 | 63.52 | 193 | 63.91 | 395 | 63.71 |
| Frequency of ever watching<br>pornographic films (n=783) *** | Never | 228 | 59.69 | 316 | 78.80 | 544 | 69.48 |
|  | 1-2 times | 101 | 26.44 | 58 | 14.46 | 159 | 20.31 |
|  | 3-4 times | 16 | 4.19 | 6 | 1.50 | 22 | 2.81 |
|  | ≥ 5 times | 37 | 9.69 | 21 | 5.24 | 58 | 7.41 |
| **Family & relationships** |  |  |  |  |  |  |  |
| Parents are alive (n=802) | None is alive | 7 | 1.79 | 2 | 0.49 | 9 | 1.12 |
| Characteristics | Categories | n | % | n | % | n | % |
|  | One parent is alive | 37 | 9.44 | 36 | 8.78 | 73 | 9.10 |
|  | Both parents are alive | 348 | 88.78 | 372 | 90.73 | 720 | 89.78 |
| Living with parents (n=733)* | None parent | 9 | 2.55 | 23 | 6.05 | 32 | 4.37 |
|  | One parent | 74 | 20.96 | 88 | 23.16 | 162 | 22.10 |
|  | Both parents | 270 | 76.49 | 269 | 70.79 | 539 | 73.53 |
| Household economic status<br>Categories (n=811)+ | Category 1/2 | 129 | 32.41 | 157 | 38.01 | 286 | 35.27 |
|  | Category 3/4 | 196 | 49.25 | 199 | 48.18 | 395 | 48.71 |
|  | Do not know | 73 | 18.34 | 57 | 13.80 | 130 | 16.03 |
| Father's level of education<br>(n=625) | Primary & lower | 123 | 41.00 | 116 | 35.69 | 239 | 38.24 |
|  | TVET | 38 | 12.67 | 37 | 11.38 | 75 | 12.00 |
|  | Secondary | 79 | 26.33 | 102 | 31.38 | 181 | 28.96 |
|  | Higher | 60 | 20.00 | 70 | 21.54 | 130 | 20.80 |
| Mother's level of education<br>(n=693) | Primary or lower | 140 | 41.67 | 165 | 46.22 | 305 | 44.01 |
|  | TVET | 31 | 9.23 | 35 | 9.80 | 66 | 9.52 |
|  | Secondary | 94 | 27.98 | 96 | 26.89 | 190 | 27.42 |
|  | Higher | 71 | 21.13 | 61 | 17.09 | 132 | 19.05 |
| Ever discussed sex matters with<br>the father (n=790) | Often | 29 | 7.53 | 24 | 5.93 | 53 | 6.71 |
|  | Occasionally | 116 | 30.13 | 102 | 25.19 | 218 | 27.59 |
|  | Never | 240 | 62.34 | 279 | 68.89 | 519 | 65.70 |
| Ever discussed sex matters with<br>the mother (n=790)*** | Often | 69 | 17.97 | 175 | 43.10 | 244 | 30.89 |
|  | Occasionally | 130 | 33.85 | 155 | 38.18 | 285 | 36.08 |
|  | Never | 185 | 48.18 | 76 | 18.72 | 261 | 33.04 |
| Ever discussed sex matters with<br>the brothers or sisters (n=788)* | Often | 37 | 9.61 | 52 | 12.90 | 89 | 11.29 |
|  | Occasionally | 94 | 24.42 | 121 | 30.02 | 215 | 27.28 |
|  | Never | 254 | 65.97 | 230 | 57.07 | 484 | 61.42 |
| Perceived ease in<br>communicating with the father | (Very) easy | 198 | 59.28 | 198 | 54.70 | 396 | 56.90 |
|  | Average | 59 | 17.66 | 69 | 19.06 | 128 | 18.39 |
| Characteristics | Categories | n | % | n | % | n | % |
| (n=696) | (very) difficult | 48 | 14.37 | 60 | 16.57 | 108 | 15.52 |
|  | Do not see him | 29 | 8.68 | 35 | 9.67 | 64 | 9.20 |
| Perceived ease in communicating with the mother (n=773) | (Very) easy | 313 | 83.47 | 324 | 81.41 | 637 | 82.41 |
|  | Average | 34 | 9.07 | 38 | 9.55 | 72 | 9.31 |
|  | (very) difficult | 22 | 5.87 | 26 | 6.53 | 48 | 6.21 |
|  | Do not see her | 6 | 1.60 | 10 | 2.51 | 16 | 2.07 |
| Perceived ease in communicating with brothers or sisters (n=789) | (Very) easy | 317 | 81.49 | 319 | 79.75 | 636 | 80.61 |
|  | Average | 36 | 9.25 | 35 | 8.75 | 71 | 9.00 |
|  | (very) difficult | 26 | 6.68 | 36 | 9.00 | 62 | 7.86 |
|  | Do not see them | 10 | 2.57 | 10 | 2.50 | 20 | 2.53 |
| **Community** |  |  |  |  |  |  |  |
| Ever accessed puberty/pregnancy information (n=811)** | Yes | 357 | 89.70 | 393 | 95.16 | 750 | 92.48 |
|  | No | 41 | 10.30 | 20 | 4.84 | 61 | 7.52 |
| Type of school (n=811) | Public | 207 | 52.01 | 217 | 52.54 | 424 | 52.28 |
|  | Private | 191 | 47.99 | 196 | 47.46 | 387 | 47.72 |
| Ever been forced to have sexual intercourse (n=750) | No | 337 | 92.08 | 353 | 91.93 | 690 | 92.00 |
|  | Yes | 29 | 7.92 | 31 | 8.07 | 60 | 8.0 |
| Ever been touched on private parts (breast, etc.) without their willingness (n=739) | No | 323 | 91.76 | 354 | 91.47 | 677 | 91.61 |
|  | Yes | 29 | 8.24 | 33 | 8.53 | 62 | 8.39 |

**Note:**
+ **Economic status categories of households:** According to Ezeanya-Esiobu (44), the Government of Rwanda categorises households into four groups according to their socioeconomic status (the Ubudehe categories): the very poor and vulnerable individuals who are unable to feed themselves (category 1), those who can access low-class accommodations, those who have no paid employment, and those who eat twice or less a day (category 2). Those who earn money from their employment belong to Category 3, and those who have permanent employment and are well-off belong to Category 4.
**TVET:** Technical and vocational education and training
\* $p \leq 0.05$ , \*\* $p \leq 0.01$ , \*\*\* $p \leq 0.001$

Concerning the perceived ease of communicating with their fathers, 396 (56.9%) indicated that this communication was easy or very easy, while 15.5% reported that it was difficult or very difficult. Although most participants indicated that they could communicate easily with their mothers (82%, n=637), 48 of them (6%) noted that this communication was difficult or very difficult. On the other hand, 637 participants (82%) expressed ease of communication with mothers, and 636 (81%) reported that it was easy or very easy to communicate with brothers or sisters.

Regarding discussing sex matters with family members, 519 (65.7%) of the 790 participants had never discussed sex matters with their fathers, while 33% had never had such a discussion with their mothers. Similarly, 484 (61%) never discussed sex issues with brothers or sisters (Table 2).

#### Community level

Seven hundred and fifty participants (92.2%) had access to SRHR information (including information on puberty and/or pregnancy), with a significantly greater percentage of girls (48.5%) than boys (44.0%) (Table 2). Most participants were from public schools (n=424, 52.3%). Sixty participants (8%) reported ever being forced to have sexual intercourse, while 62 (8.4%) participants indicated ever being touched on private parts against their will.

### Early adolescents’ sexual experience

#### Nonpenetrative sexual experiences

In terms of nonpenetrative sexual experience, participants reported having held hands (73.6%, n=787), hugged (58.6%, n=783), kissed their lips (13.3%, n=769), or caressed each other’s body (12.1%, n=780). Twenty-three of the 757 participants (3%) indicated that they had ever stroked their partner’s genitals to climax, and 20 of the 779 participants (2.6%) indicated that their partners had done this to them. Touching the partner’s genitals was reported by 34 of 773 (4.4%) participants, while 36 of 764 participants (4.7%) indicated that their partners had ever touched theirs. Overall, the findings show that 532 of the 658 participants (80.9%) reported some form of nonpenetrative sexual experience (Table 3). In bivariate analysis, the factors associated with nonpenetrative sexual experience without adjustment for other factors included ever using mobile phones (COR= 1.6, 95% CI= 1.1-2.4); ever attending parties (COR= 9.9, 95% CI= 2.4-41.1); ever drinking alcohol (COR= 4.1, 95% CI= 1.6-10.3); ever using the internet (COR= 1.7, 95% CI= 1.1-2.5); and their father having a primary or lower education level (COR= 1.9, 95% CI= 1.1-3.5). Multivariate logistic regression analysis revealed that ever attending parties where young people danced (AOR= 6.8, 95% CI= 1.6-29.2), ever using the internet (AOR= 1.9, 95% CI= 1.1-3.3), and having a father with lower education level (primary or lower: AOR=2.6, 95% CI= 1.3-5.0, and secondary education: AOR=1.9, 95%CI: 1.0-3.8) were factors significantly associated with nonpenetrative sexual experience (Table 4).

**Table 3:** Sexual experience of early adolescents by biological sex.

| Sexual experience | Gender |  |  |  |  |  |  |  |
| --- | --- | --- | --- | --- | --- | --- | --- | --- |
|  |  | Male |  | Female |  | All |  | p-value |
|  |  | n | % | n | % | n | % |  |
| Non-penetrative experience |  |  |  |  |  |  |  |  |
| Hugging | Yes | 231 | 60.2 | 228 | 57.1 | 459 | 58.6 | 0.392 |
|  | No | 153 | 39.8 | 171 | 42.9 | 324 | 41.4 |  |
|  | Total | 384 | 100.0 | 399 | 100.0 | 783 | 100.0 |  |
| Holding hands | Yes | 290 | 74.9 | 289 | 72.3 | 579 | 73.6 | 0.393 |
|  | No | 97 | 25.1 | 111 | 27.8 | 208 | 26.4 |  |
|  | Total | 387 | 100.0 | 400 | 100.0 | 787 | 100.0 |  |
| Caressing each other's bodies | Yes | 63 | 16.4 | 31 | 7.8 | 94 | 12.1 | <0.001 |
|  | No | 321 | 83.6 | 365 | 92.2 | 686 | 87.9 |  |
|  | Total | 384 | 100.0 | 396 | 100.0 | 780 | 100.0 |  |
| Kissing a boy/girlfriend/partner on the lips | Yes | 61 | 15.9 | 41 | 10.6 | 102 | 13.3 | 0.030 |
|  | No | 322 | 84.1 | 345 | 89.4 | 667 | 86.7 |  |
|  | Total | 383 | 100.0 | 386 | 100.0 | 769 | 100.0 |  |
| Touching a boy/girlfriend/partner's vagina/penis with a hand | Yes | 27 | 7.1 | 7 | 1.8 | 34 | 4.4 | <0.001 |
|  | No | 354 | 92.9 | 385 | 98.2 | 739 | 95.6 |  |
|  | Total | 381 | 100.0 | 392 | 100.0 | 773 | 100.0 |  |
| Stroking a boy/girlfriend/partner's vagina/penis so that she/he climaxed | Yes | 18 | 4.9 | 5 | 1.3 | 23 | 3.0 | 0.004 |
|  | No | 348 | 95.1 | 386 | 98.7 | 734 | 97.0 |  |
|  | Total | 366 | 100.0 | 391 | 100.0 | 757 | 100.0 |  |
| His/her boy/girlfriend/partner ever touch her/his penis/vagina with her/his hand | Yes | 26 | 6.8 | 10 | 2.6 | 36 | 4.7 | 0.006 |
|  | No | 354 | 93.2 | 374 | 97.4 | 728 | 95.3 |  |
|  | Total | 380 | 100.0 | 384 | 100.0 | 764 | 100.0 |  |
| His/her boy/girlfriend/partner ever stroke his/her penis/vagina so that you climaxed | Yes | 16 | 4.2 | 4 | 1.0 | 20 | 2.6 | 0.006 |
|  | No | 369 | 95.8 | 390 | 99.0 | 759 | 97.4 |  |
|  | Total | 385 | 100.0 | 394 | 100.0 | 779 | 100.0 |  |
| Prevalence of nonpenetrative sexual experience | Yes | 262 | 81.9 | 270 | 79.9 | 532 | 80.9 | 0.516 |
|  | No | 58 | 18.1 | 68 | 20.1 | 126 | 19.1 |  |
|  | Total | 320 | 100.0 | 338 | 100.0 | 658 | 100.0 |  |
| **Penetrative experience** |  |  |  |  |  |  |  |  |
| Ever had oral sex | Yes | 18 | 4.6 | 3 | 0.7 | 21 | 2.6 | 0.001 |
|  | No | 373 | 95.4 | 402 | 99.3 | 775 | 97.4 |  |
|  | Total | 391 | 100.0 | 405 | 100.0 | 796 | 100.0 |  |
| Ever had anal intercourse | Yes | 21 | 5.4 | 1 | 0.2 | 22 | 2.8 | <0.001 |
|  | No | 369 | 94.6 | 402 | 99.8 | 771 | 97.2 |  |
|  | Total | 390 | 100.0 | 403 | 100.0 | 793 | 100.0 |  |
| Ever had vaginal intercourse with | Yes | 26 | 6.9 | 10 | 2.5 | 36 | 4.7 | 0.004 |
| anyone | No | 353 | 93.1 | 384 | 97.5 | 737 | 95.3 |  |
|  | Total | 379 | 100.0 | 394 | 100.0 | 773 | 100.0 |  |
| Prevalence of penetrative sexual | Yes | 41 | 11.1 | 12 | 3.1 | 53 | 7.0 | <0.001 |
| experience | No | 330 | 88.9 | 376 | 96.9 | 706 | 93.0 |  |
|  | Total | 371 | 100.0 | 388 | 100.0 | 759 | 100.0 |  |

**Table 4:**
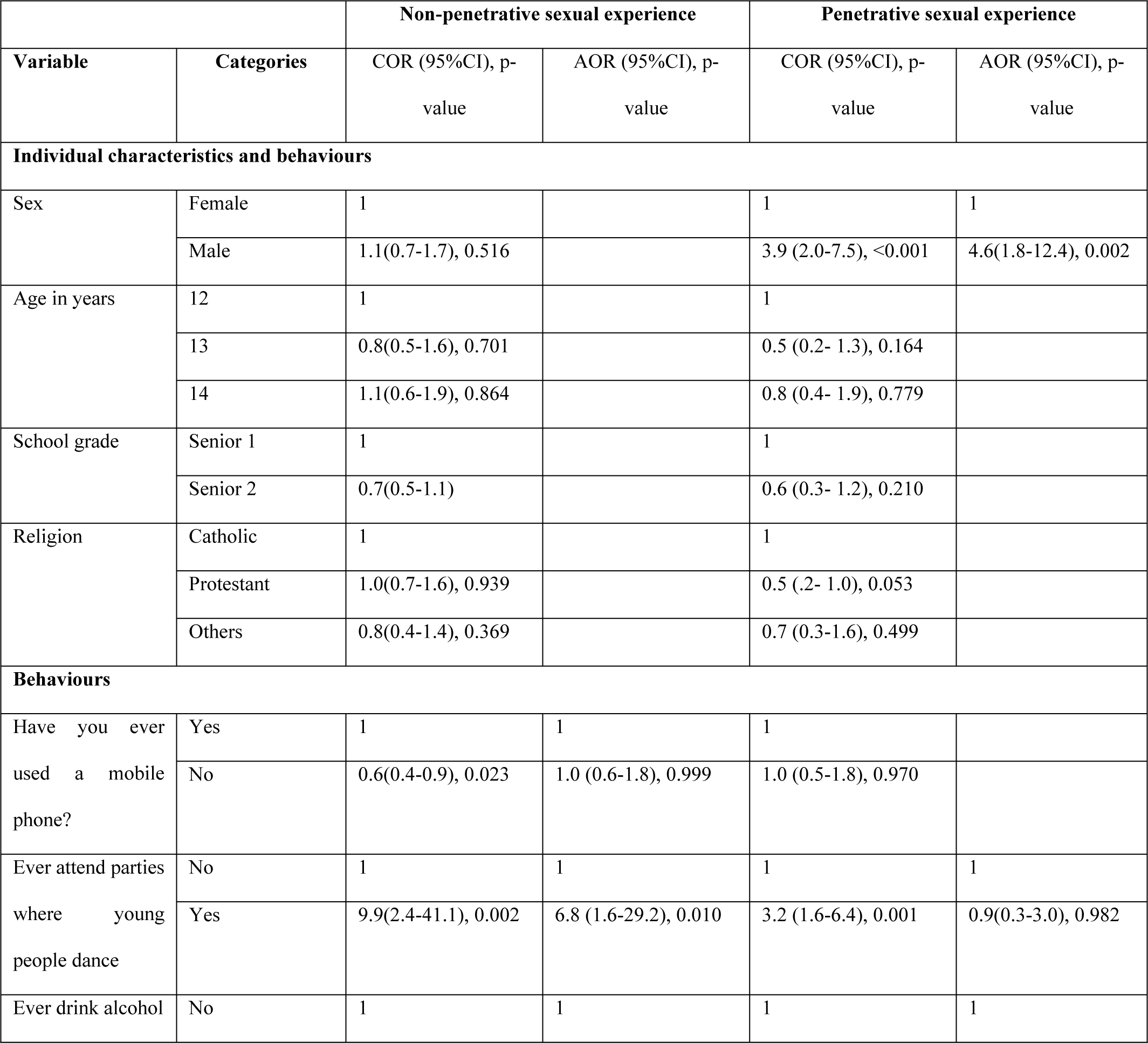
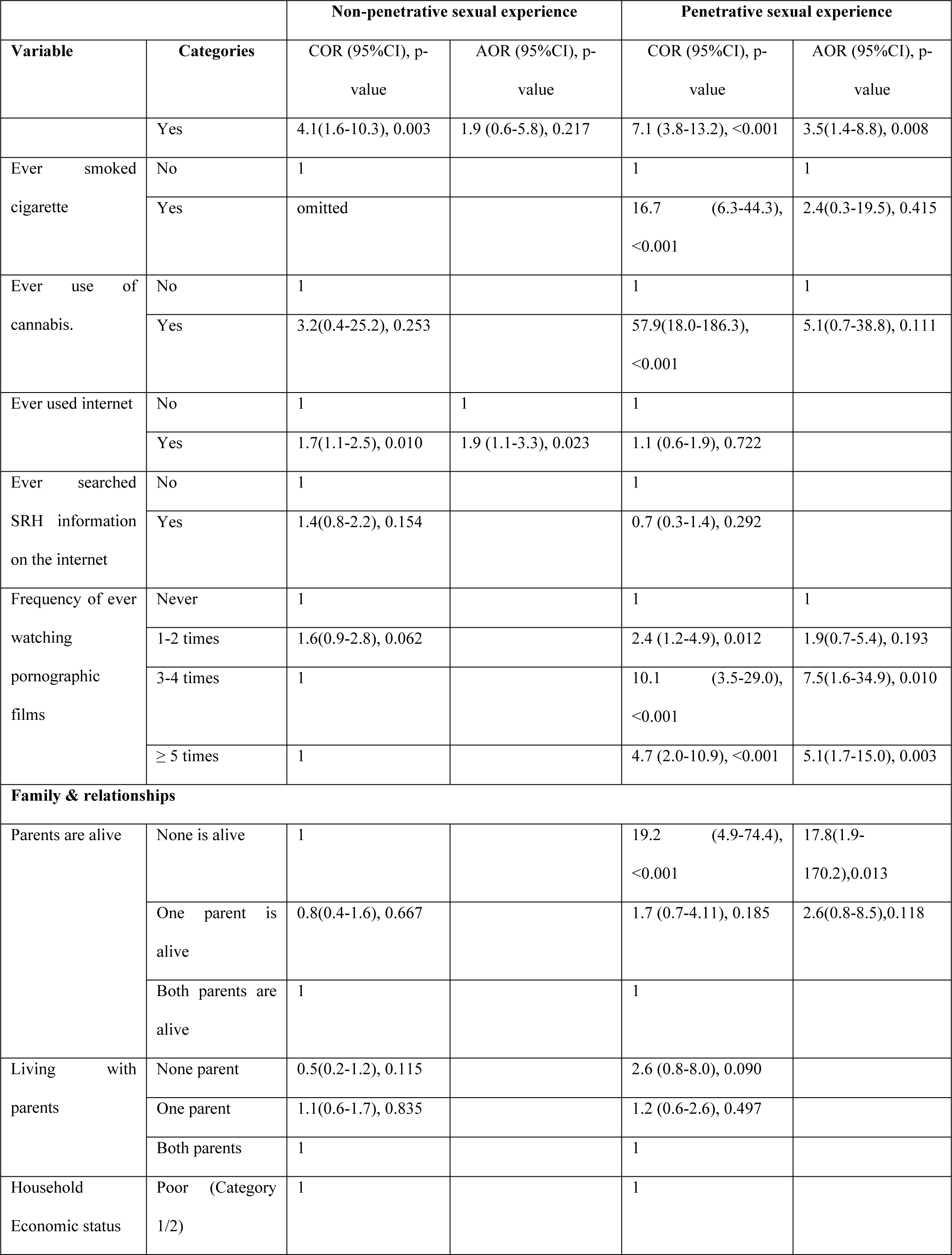
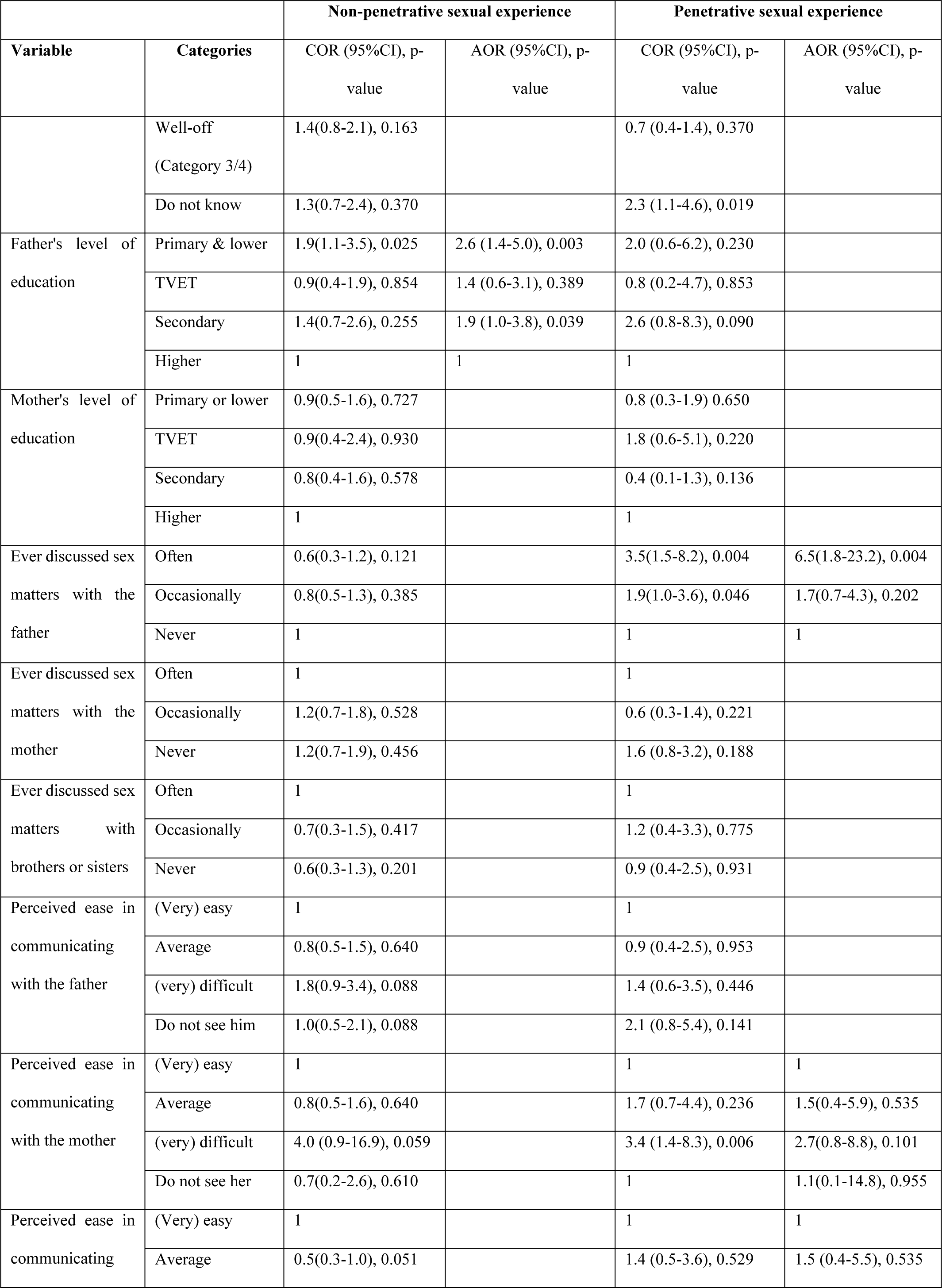
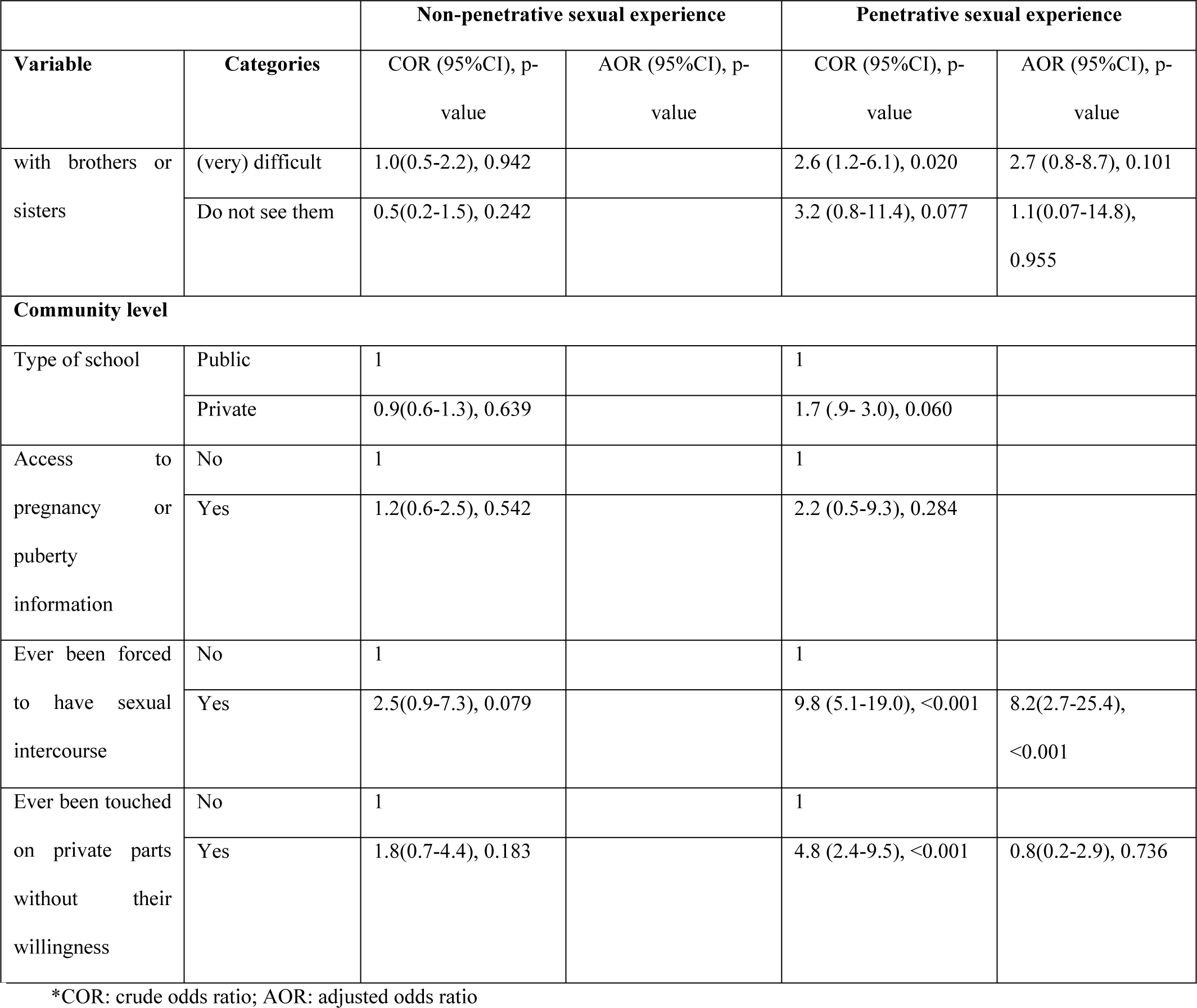
Results from the bivariate and multivariate logistic regression analyses between participants’ sexual experience and social-ecological covariates.

|  |  | Non-penetrative sexual experience |  | Penetrative sexual experience |  |
| --- | --- | --- | --- | --- | --- |
| Variable | Categories | COR (95%CI), p-value | AOR (95%CI), p-value | COR (95%CI), p-value | AOR (95%CI), p-value |
| **Individual characteristics and behaviours** |  |  |  |  |  |
| Sex | Female | 1 |  | 1 | 1 |
|  | Male | 1.1(0.7-1.7), 0.516 |  | 3.9 (2.0-7.5), <0.001 | 4.6(1.8-12.4), 0.002 |
| Age in years | 12 | 1 |  | 1 |  |
|  | 13 | 0.8(0.5-1.6), 0.701 |  | 0.5 (0.2- 1.3), 0.164 |  |
|  | 14 | 1.1(0.6-1.9), 0.864 |  | 0.8 (0.4- 1.9), 0.779 |  |
| School grade | Senior 1 | 1 |  | 1 |  |
|  | Senior 2 | 0.7(0.5-1.1) |  | 0.6 (0.3- 1.2), 0.210 |  |
| Religion | Catholic | 1 |  | 1 |  |
|  | Protestant | 1.0(0.7-1.6), 0.939 |  | 0.5 (.2- 1.0), 0.053 |  |
|  | Others | 0.8(0.4-1.4), 0.369 |  | 0.7 (0.3-1.6), 0.499 |  |
| **Behaviours** |  |  |  |  |  |
| Have you ever used a mobile phone? | Yes | 1 | 1 | 1 |  |
|  | No | 0.6(0.4-0.9), 0.023 | 1.0 (0.6-1.8), 0.999 | 1.0 (0.5-1.8), 0.970 |  |
| Ever attend parties where young people dance | No | 1 | 1 | 1 | 1 |
|  | Yes | 9.9(2.4-41.1), 0.002 | 6.8 (1.6-29.2), 0.010 | 3.2 (1.6-6.4), 0.001 | 0.9(0.3-3.0), 0.982 |
| Ever drink alcohol | No | 1 | 1 | 1 | 1 |
| Variable | Categories | COR (95%CI), p-value | AOR (95%CI), p-value | COR (95%CI), p-value | AOR (95%CI), p-value |
|  | Yes | 4.1(1.6-10.3), 0.003 | 1.9 (0.6-5.8), 0.217 | 7.1 (3.8-13.2), <0.001 | 3.5(1.4-8.8), 0.008 |
| Ever smoked cigarette | No | 1 |  | 1 | 1 |
|  | Yes | omitted |  | 16.7 (6.3-44.3), <0.001 | 2.4(0.3-19.5), 0.415 |
| Ever use of cannabis. | No | 1 |  | 1 | 1 |
|  | Yes | 3.2(0.4-25.2), 0.253 |  | 57.9(18.0-186.3), <0.001 | 5.1(0.7-38.8), 0.111 |
| Ever used internet | No | 1 | 1 | 1 |  |
|  | Yes | 1.7(1.1-2.5), 0.010 | 1.9 (1.1-3.3), 0.023 | 1.1 (0.6-1.9), 0.722 |  |
| Ever searched SRH information on the internet | No | 1 |  | 1 |  |
|  | Yes | 1.4(0.8-2.2), 0.154 |  | 0.7 (0.3-1.4), 0.292 |  |
| Frequency of ever watching pornographic films | Never | 1 |  | 1 | 1 |
|  | 1-2 times | 1.6(0.9-2.8), 0.062 |  | 2.4 (1.2-4.9), 0.012 | 1.9(0.7-5.4), 0.193 |
|  | 3-4 times | 1 |  | 10.1 (3.5-29.0), <0.001 | 7.5(1.6-34.9), 0.010 |
|  | ≥ 5 times | 1 |  | 4.7 (2.0-10.9), <0.001 | 5.1(1.7-15.0), 0.003 |
| **Family & relationships** |  |  |  |  |  |
| Parents are alive | None is alive | 1 |  | 19.2 (4.9-74.4), <0.001 | 17.8(1.9-170.2),0.013 |
|  | One parent is alive | 0.8(0.4-1.6), 0.667 |  | 1.7 (0.7-4.11), 0.185 | 2.6(0.8-8.5),0.118 |
|  | Both parents are alive | 1 |  | 1 |  |
| Living with parents | None parent | 0.5(0.2-1.2), 0.115 |  | 2.6 (0.8-8.0), 0.090 |  |
|  | One parent | 1.1(0.6-1.7), 0.835 |  | 1.2 (0.6-2.6), 0.497 |  |
|  | Both parents | 1 |  | 1 |  |
| Household Economic status | Poor (Category 1/2) | 1 |  | 1 |  |
| Variable | Categories | COR (95%CI), p-value | AOR (95%CI), p-value | COR (95%CI), p-value | AOR (95%CI), p-value |
|  | Well-off<br>(Category 3/4) | 1.4(0.8-2.1), 0.163 |  | 0.7 (0.4-1.4), 0.370 |  |
|  | Do not know | 1.3(0.7-2.4), 0.370 |  | 2.3 (1.1-4.6), 0.019 |  |
| Father's level of education | Primary & lower | 1.9(1.1-3.5), 0.025 | 2.6 (1.4-5.0), 0.003 | 2.0 (0.6-6.2), 0.230 |  |
|  | TVET | 0.9(0.4-1.9), 0.854 | 1.4 (0.6-3.1), 0.389 | 0.8 (0.2-4.7), 0.853 |  |
|  | Secondary | 1.4(0.7-2.6), 0.255 | 1.9 (1.0-3.8), 0.039 | 2.6 (0.8-8.3), 0.090 |  |
|  | Higher | 1 | 1 | 1 |  |
| Mother's level of education | Primary or lower | 0.9(0.5-1.6), 0.727 |  | 0.8 (0.3-1.9) 0.650 |  |
|  | TVET | 0.9(0.4-2.4), 0.930 |  | 1.8 (0.6-5.1), 0.220 |  |
|  | Secondary | 0.8(0.4-1.6), 0.578 |  | 0.4 (0.1-1.3), 0.136 |  |
|  | Higher | 1 |  | 1 |  |
| Ever discussed sex matters with the father | Often | 0.6(0.3-1.2), 0.121 |  | 3.5(1.5-8.2), 0.004 | 6.5(1.8-23.2), 0.004 |
|  | Occasionally | 0.8(0.5-1.3), 0.385 |  | 1.9(1.0-3.6), 0.046 | 1.7(0.7-4.3), 0.202 |
|  | Never | 1 |  | 1 | 1 |
| Ever discussed sex matters with the mother | Often | 1 |  | 1 |  |
|  | Occasionally | 1.2(0.7-1.8), 0.528 |  | 0.6 (0.3-1.4), 0.221 |  |
|  | Never | 1.2(0.7-1.9), 0.456 |  | 1.6 (0.8-3.2), 0.188 |  |
| Ever discussed sex matters with brothers or sisters | Often | 1 |  | 1 |  |
|  | Occasionally | 0.7(0.3-1.5), 0.417 |  | 1.2 (0.4-3.3), 0.775 |  |
|  | Never | 0.6(0.3-1.3), 0.201 |  | 0.9 (0.4-2.5), 0.931 |  |
| Perceived ease in communicating with the father | (Very) easy | 1 |  | 1 |  |
|  | Average | 0.8(0.5-1.5), 0.640 |  | 0.9 (0.4-2.5), 0.953 |  |
|  | (very) difficult | 1.8(0.9-3.4), 0.088 |  | 1.4 (0.6-3.5), 0.446 |  |
|  | Do not see him | 1.0(0.5-2.1), 0.088 |  | 2.1 (0.8-5.4), 0.141 |  |
| Perceived ease in communicating with the mother | (Very) easy | 1 |  | 1 | 1 |
|  | Average | 0.8(0.5-1.6), 0.640 |  | 1.7 (0.7-4.4), 0.236 | 1.5(0.4-5.9), 0.535 |
|  | (very) difficult | 4.0 (0.9-16.9), 0.059 |  | 3.4 (1.4-8.3), 0.006 | 2.7(0.8-8.8), 0.101 |
|  | Do not see her | 0.7(0.2-2.6), 0.610 |  | 1 | 1.1(0.1-14.8), 0.955 |
| Perceived ease in communicating | (Very) easy | 1 |  | 1 | 1 |
|  | Average | 0.5(0.3-1.0), 0.051 |  | 1.4 (0.5-3.6), 0.529 | 1.5 (0.4-5.5), 0.535 |
| Variable | Categories | COR (95%CI), p-value | AOR (95%CI), p-value | COR (95%CI), p-value | AOR (95%CI), p-value |
| with brothers or sisters | (very) difficult | 1.0(0.5-2.2), 0.942 |  | 2.6 (1.2-6.1), 0.020 | 2.7 (0.8-8.7), 0.101 |
|  | Do not see them | 0.5(0.2-1.5), 0.242 |  | 3.2 (0.8-11.4), 0.077 | 1.1(0.07-14.8), 0.955 |
| **Community level** |  |  |  |  |  |
| Type of school | Public | 1 |  | 1 |  |
|  | Private | 0.9(0.6-1.3), 0.639 |  | 1.7 (.9- 3.0), 0.060 |  |
| Access to pregnancy or puberty information | No | 1 |  | 1 |  |
|  | Yes | 1.2(0.6-2.5), 0.542 |  | 2.2 (0.5-9.3), 0.284 |  |
| Ever been forced to have sexual intercourse | No | 1 |  | 1 |  |
|  | Yes | 2.5(0.9-7.3), 0.079 |  | 9.8 (5.1-19.0), <0.001 | 8.2(2.7-25.4), <0.001 |
| Ever been touched on private parts without their willingness | No | 1 |  | 1 |  |
|  | Yes | 1.8(0.7-4.4), 0.183 |  | 4.8 (2.4-9.5), <0.001 | 0.8(0.2-2.9), 0.736 |

\*COR: crude odds ratio; AOR: adjusted odds ratio

#### Penetrative experience

The results highlighted that some participants had experienced some form of penetrative sex, including vaginal, anal, or oral sex. The findings indicated that 36 of 773 participants (4.6%) have had vaginal intercourse, 22 of the 793 (2.7%) have had anal intercourse, and 21 of the 796 participants (2.6%) reported ever having had oral sex. Overall, 53 of 759 (7.0%) reported any of these activities involving penetrative sexual experience (oral, vaginal, anal penetration) (Table 3). The bivariate analysis of penetrative sexual experience indicated that the factors associated with penetrative sexual experience included male sex (COR= 3.9, 95% CI= 2.0-7.5), ever attending parties where young people danced (COR= 3.2, 95% CI= 1.6-6.4), ever drinking alcohol (COR= 7.1, 95% CI= 3.8-13.2), ever smoking cigarettes (COR= 16.7, 95% CI= 6.3-44.3), ever using cannabis (COR=57.9, 95% CI= 18.0-186.3), and ever watching pornographic films (COR= 3.5, 95% CI= 1.9-6.4). In addition, participants whose both parents were not alive (COR= 19.2, 95% CI= 4.9-74.4), who perceived that communication with their mothers was difficult (COR= 3.4, 95% CI= 1.4-8.3), and whose communication with brothers or sisters was perceived as difficult (COR= 2.6, 95% CI= 1.2-6.1) had higher odds of penetrative sexual experience. Additional factors associated with penetrative sexual experience in the bivariate analysis included ever being forced to have sexual intercourse (COR= 9.8, 95% CI= 5.1-19.0) and ever being touched on private parts (COR= 4.8, 95% CI= 2.4-9.5). Discussing sex matters often with fathers was associated with higher odds of penetrative sexual experience (COR= 3.5, 95% CI=1.5-8.2). The multivariate logistic regression analysis indicated that male participants (AOR= 4.6, 95% CI= 1-8-12.4) ever drinking alcohol (AOR= 3.5, 95% CI= 1.4-8.8), ever watching pornography (3-5 times, AOR= 7.5, 95% CI= 1.6-34.9; ≥ five times, (AOR= 5.1, 95% CI= 1.7-15.0); being full orphan (AOR= 17.8, 95% CI= 1.9-170.2), and ever being forced to have sexual intercourse (AOR= 8.2, 95% CI= 2.7-25.4). In addition, often discussing sex matters with one’s father (AOR= 6.5, 95% CI= 1.8-23.2) was associated with higher odds of penetrative sexual experience (Table 4).

## Discussion

This study aimed to assess the sexual experiences and the social-ecological factors associated with such experiences among early adolescents from selected districts in Rwanda. The findings indicated that more than four out of five early adolescents (81%) had started engaging in at least one nonpenetrative sexual activity, including hugging, holding hands, caressing, and touching or being touched on their genitals. Concerning being touched by genitals among girls, the prevalence ranged from 1 to 2.6% in the present study, which corroborates the findings of Maina et al., who reported a prevalence of 1% among very young adolescent girls in Nairobi, Kenya [45]. However, the prevalence of caressing in this study was 7.8% among girls, which Maina et al. [45] did not include. Furthermore, this study found a greater prevalence of hugging (57.1%) and holding hands (72.3%) than was found by Maina et al., which were respectively 6% and 9% among very young adolescent girls in Nairobi, Kenya [45]. In addition, the prevalence of kissing was greater among girls (10.6%) in the present study, while Maina et al. reported 2% of young adolescents in Nairobi, Kenya [45]. In a different study in Kenya, Kågesten et al. indicated that most early adolescents have not engaged in romantic and sexual activities [23]. Findings from early adolescents (11-14 years) in Korogocho slums, Kenya, indicated that the proportions of hugging (8.6%), kissing (2.7%), holding hands (10.5%), and fondling (2.5%) were lower than those in this study, with boys having higher proportions than girls [23]. In general, these findings suggest that early adolescents experience some nonpenetrative sexual activities, with differences among boys and girls.

Concerning the factors associated with nonpenetrative sexual experience, logistic regression analysis revealed that attending parties involving other young people, using the internet, and having a primary and secondary education level of the participants’ fathers were found to be independent predictors of nonpenetrative sexual experiences. The literature highlights the role of the internet in the provision of SRHR information [46–50] and in influencing the sexual behaviours of adolescents. For example, pornography and sexually explicit photographs teach young adolescents about sex and what it looks like, which may have both positive and negative consequences for the sexual behaviours of adolescents [50, 51]. Parents are the main sources of SRHR information for adolescents [15, 52]. They constitute important models in their children’s lives and transmit values, traditions, and lifestyles that inform healthy decision-making about sexual health [53]. Furthermore, improved communication between parents and adolescents contributes to healthy sexual behaviours [22, 54, 55], and parents’ education may be beneficial in promoting adolescent reproductive health [56]. However, discussing sex matters often with one’s father was a risk factor for penetrative sexual experience in this study from penetrative sexual experience. This finding contrasts with the knowledge that parent-child communication about sexual issues enhances healthy sexual behaviours among adolescents [22, 57–59].

Nearly 7% of all early adolescents, including 11% of boys and 3% of girls, reported penetrative sexual experiences, including oral sex and anal and/or vaginal intercourse. Studies of early adolescents (12-15 years) have shown similar estimates (6.9%) of lifetime sexual intercourse in Low and Middle-income Countries (LMICs) [60] and among American Indian and Alaskan Native Youth aged 12-14 years (6.5%) [35]. Similarly, Roman et al. reported that sexual initiation among Brazilian early adolescents (12-14 years) was lower in girls (7%) than in boys (18%) [61], which was similar to the trend observed in this study and others in LMICs and Korea [60, 62]. Nevertheless, penetrative sexual experience was slightly lower in Indonesia (5.3%, including 6.9% of boys and 3.8% of girls) than in the present study, as indicated in the 2015 Global School-based Health Survey by Rizkianti et al. [63]. In addition, Maina et al. reported that the proportion of girls (10-14 years) with sexual intercourse was 2% in Nairobi, Kenya [45], compared to 3% in this study. The prevalence of sexual intercourse before age 13 in the United States of America was found to be slightly greater than that in the present study (7.6%) [64]. In contrast, Magnusson reported that 17% of adolescents were younger than 15 years at first sexual intercourse [65], and Hellerstedt et al. reported that 42% of American Indian Youths aged 13-15 years had ever had sexual intercourse [66]. In addition, Kushal et al. reported that early sexual initiation was 14.2% among early adolescents (12-15 years) in 50 countries where the Global School-based Health Survey was conducted between 2009 and 2015. The prevalence of penetrative sexual experiences among early adolescents is in line with the findings of other studies in LMICs. Nonetheless, this prevalence differs in the United States of America, where a higher prevalence has been reported, as well as in Indonesia and Kenya, where lower penetrative sexual experiences have been indicated.

Concerning the factors associated with penetrative sexual experience, this study showed that ever being forced to have sexual intercourse was strongly associated with penetrative sex. On this point, the literature indicates that early sexual experience among adolescents occurs in the context of sexual coercion and violence [1, 13, 67]. This study also revealed that a high frequency of watching pornographic films was associated with penetrative sexual experience, which is consistent with previous studies indicating that exposure to pornography may have harmful effects on sexual maturation and behaviours in adolescents [17, 68, 69] and that first sex was associated with exposure to pornography [70–72]. In addition, another study involving adolescents aged 10-19 years in rural eastern Uganda found that watching sexually explicit films was associated with ever having sexual intercourse [73]. Furthermore, pornography use by adolescents results in unprotected penetrative and oral sex, a high frequency of casual sex, and an earlier onset of sexual activity [74]. The current study also highlighted that male participants were more likely to experience penetrative sexual experiences, which is consistent with findings from Reis et al. [75], Kushal et al. [76], and Roman et al. [61]. This may be due to masculinity norms encouraging boys to early sexual initiation [75].

Smoking and ever-drinking alcohol in the present study were associated with penetrative sexual experiences. This finding is consistent with other research indicating that smoking cigarettes and using alcohol and other drugs were associated with sexual intercourse among in-school adolescents from five sub-Saharan African countries [77] and among older adolescents and youths [78]. Furthermore, a study of the factors associated with sexual initiation among participants, including early adolescents in Brazil, revealed that the use of alcoholic beverages was associated with early sexual initiation [61, 79]. Similarly, a higher prevalence of early sexual initiation (14.2%) was reported in another study involving early adolescents aged 12-15 years in 50 countries [76].

### Study limitations and strengths

This study has several limitations. One of them is the use of cross-sectional data to determine causality. Another limitation of this study is the generalizability of the findings. However, the selected number of participants was representative of the study population, and their data provided insights into the SRHR knowledge and sexual experience of early adolescents in Rwanda. In addition, the findings were obtained based on participants’ self-reports about sexuality, which might have been affected by social desirability, especially when reporting (underreporting or overreporting) sexual experiences [80, 81]. We minimised this through privacy, anonymity and voluntary participation. Finally, there is the possibility of recall bias because participants had to remember and report their sexual experiences, which suggests that they might have forgotten important information related to their sexual experiences. Despite these limitations, this study contributes to the extant body of literature on early adolescents by providing an overview of the prevalence and factors associated with sexual experience in sub-Saharan Africa, specifically in Rwanda. This study enriches the body of literature and provides baseline information for SRHR interventions to improve the sexual and reproductive health and rights of early adolescents in Rwanda and elsewhere.

## Conclusion

This study showed that the prevalence of nonpenetrative sexual experience among early adolescents was 81%, and 7% of the participants reported penetrative sexual experience. Penetrative sexual experience at the individual level was associated with being male se, and behaviours such as drinking alcohol and watching films showing sexual content. Being a double orphan and often discussing sex matters with one’s father were independently associated with increased odds of penetrative sexual experience (family and relationships level). Being forced to have sexual intercourse was also associated with penetrative sexual experience (community level). The study showed that behaviours such as attending parties, using the internet, and reporting a low father’s education level (family level) were associated with nonpenetrative sexual experiences. These findings suggest the need for public health interventions, including educating early adolescents, especially boys, about SRHR, reducing alcohol consumption, addressing child abuse, monitoring access to SRHR information sources, especially the internet, scaling up parental education levels, especially about SRHR, and discussing SRHR with family members. These findings have implications for achieving sustainable development goals (SDGs) related to early adolescent sexual and reproductive health and rights in Rwanda, regionally and globally.

## Data Availability

All relevant data are within the manuscript.

## Acknowledgements

This research was supported by the Consortium for Advanced Research Training in Africa (CARTA). CARTA is jointly led by the African Population and Health Research Center and the University of the Witwatersrand and funded by the Carnegie Corporation of New York (Grant No. G-19-57145), Sida (Grant No:54100113), Uppsala Monitoring Center, Norwegian Agency for Development Cooperation (Norad), and by the Wellcome Trust [reference no. 107768/Z/15/Z] and the UK Foreign, Commonwealth & Development Office, with support from the Developing Excellence in Leadership, Training, and Science in Africa (DELTAS Africa) programme. The statements made and views expressed are solely the responsibility of the Fellow.

